# Modified FRAX Score for Prediction of Fall-induced Hip Fractures; The Added Value of Fall Energy, Number, and Social Vulnerability Index

**DOI:** 10.1101/2024.01.27.24301867

**Authors:** Atta Taseh, Evan Sirls, George Casey, Sarah Hearns, Job N. Doornberg, Santiago A. Lozano-Calderon, Mitchel B Harris, Soheil Ashkani-Esfahani

## Abstract

**Background:** The Fracture Risk Assessment Tool (FRAX), widely used for predicting the 10-year likelihood of hip fractures, does not incorporate factors like prior falls and sociodemographic characteristics, notably the Social Vulnerability Index (SVI). Recognizing these limitations, we aim to evaluate the predictive accuracy of FRAX by integrating fall frequency, fall energy, and SVI into the model for assessing the risk of fall-induced hip fractures.

**Methods:** A retrospective case-control study was conducted, and patients aged ≥ 40 years with a documented diagnosis of a fall-induced hip fracture were age-matched with controls with a history of falls without an associated hip fracture. Basic demographic data, along with information about the number of prior falls and the energy of the current falls, were collected. The FRAX and SVI were calculated accordingly. Logistic regression analysis was employed to identify significant predictors. The performance of the models was evaluated and reported using appropriate metrics. Baseline characteristics of the dataset were presented as medians with interquartile ranges (IQR) or as percentages, where applicable. The significance of the identified variables was quantified using Odds Ratio (OR) along with their 95% Confidence Interval (CI). A p-value threshold of 0.05 was set for statistical significance.

**Results:** A total of 261 patients per group were included with a median age of 74 (IQR 67-80) and 72 (IQR 62-83) years. The FRAX score was significantly associated with the likelihood of experiencing a fall-induced hip fracture, as indicated by an OR of 1.06 (CI: 1.03-1.09). Participants with a one-time history of falls had an OR of 1.58 (CI: 1.02-2.37), compared to 1.84 (CI: 1.09-3.1) for those with multiple falls. The white race, along with the Housing Type and Transportation domain of the SVI, also demonstrated to play a role (OR= 2.85 (CI: 1.56-5.2) and OR= 0.3 (CI: 0.12-0.8), respectively).

**Conclusion:** This study underscored the significance of factors such as fall frequency, SVI, and race in predicting fall-induced hip fractures. It also highlighted the need for further refinement of the FRAX tool. We recommend that future research should be focused on validating the impact of these sociodemographic and fall characteristics on a broader scale, along with exploring the implications of clinical surrogates related to falls.

## Introduction

Hip fractures are a debilitating health concern worldwide that is estimated to rise from 1.26 million cases globally in 1990 to 4.5 million cases by 2050.^1, 2^ An estimated 37.3 million falls occur annually, contributing to over 95% of hip fractures.^1,3,4^ According to the literature, a total of 1.75 million disability-adjusted life years (DALYs) are lost globally, accounting for 0.1% of the worldwide disease burden and 1.4% of the disease burden among women in established market economies.^5^ Therefore, predicting and implementing early intervention strategies is of crucial value to decrease the burden of hip fractures associated with falls.

To accomplish this goal, the Fracture Risk Assessment Tool (FRAX) generated by the World Health Organization (WHO) has been developed to predict the 10-year probability of an adult having a hip fracture.^6^ FRAX utilizes the most important risk factors known for major osteoporotic fractures, including age, sex, weight, height, alcohol use, glucocorticoid use, femoral neck bone mineral density (BMD), history of previous fracture in the patients and their parents, smoking status, and rheumatoid arthritis.^7^ With 6 million yearly calculations done in 173 countries, FRAX has been immensely helpful in predicting fracture risks. However, it is frequently criticized for lacking a detailed and standardized description of each risk factor, including the recency of fracture and glucocorticoid dose.^8^ Furthermore, since falls are one of the most critical factors leading to hip fractures in older adults, a common criticism of FRAX is that it does not factor in fall history.^8, 9^

Since the official launch of FRAX in 2008, concerns have arisen regarding its inability to consider the impact of falls.^10,11^ In 2010, the Task Force acknowledged this concern, stating that “Fracture probability may be underestimated in individuals with a history of frequent falls, but quantification of this risk is not currently possible.” The primary reason for this challenge was the lack of sufficient evidence available at that time. Since then, the body of literature on this topic has expanded, with controversial results suggesting a complex relationship between FRAX and falls and highlighting the need for a more careful approach when integrating the two.^12–14^ Furthermore, while much of the research focuses on this aspect, other fall characteristics seem not well investigated. Among these, the mechanics of the falls seem to play a significant role in the risk of hip fractures.^15^ Hayes et al. pointed out that the energy or impact of the fall is found to be a crucial factor when assessing the risk factors for hip fractures.^16^ This suggests that not just the frequency of falls but also their nature and severity might be essential factors in assessing fall-induced hip fracture risk.

Given the multifactorial nature of hip fractures, efforts have been made to point out the relevant demographic and other non-medical risk factors in hip fractures.^17–20^ Some factors, including age and sex, are already addressed in the FRAX, while others, like race, ethnicity, and socioeconomic factors, are not. The Social Vulnerability Index (SVI), developed by the Centers for Disease Control and Prevention (CDC), is intended to measure the vulnerability of communities to environmental hazards, disease outbreaks, and other public health emergencies.^21^ It encompasses various subscales, including Socioeconomic Status, Household Characteristics, Racial and Ethnic Minority Status, Housing Type and Transportation. When used in medicine, it enables capturing the pre-existing social inadequacies that make individuals or communities disproportionately vulnerable to a particular medical condition or disease.^22^ By understanding these correlations, healthcare providers and policymakers can develop more targeted interventions and resource allocations to reduce the risk of fall-induced injuries in the most vulnerable populations.

In this study, we hypothesized that fall frequency and energy, along with the SVI, are of predictive value in fall-induced hip fractures. Moreover, in order to have a more comprehensive and accurate assessment of hip fracture risk, we aim to develop a modified version of the FRAX that incorporates additional variables found to be significant risk factors for fall-induced hip fractures in our study.

## Methods and Materials

### I. Study Design and Population

We conducted a retrospective case-control study through a tertiary institution located in Boston, Massachusetts. After getting approval from the institutional review board (IRB # 2023P000741), data were queried through the institution’s data repository, the Research Patient Data Registry (RPDR). The relevant ICD-9, ICD-10, and CPT codes, along with string searches for clinician notes, were used for the query. The initial results were screened, and participants aged ≥ 40 years with a confirmed diagnosis of fall-induced hip fracture were included as cases and those who experienced falls without a resultant hip injury as controls. A total of 2,132 patients with a history of falls who were assessed for hip fractures were screened. Cases (patients with hip fracture) and controls (without hip fracture) were age-matched for the final analysis. The study excluded individuals who did not have a mention of the mechanism of falls on their charts or experienced falls resulting from violent encounters and animal attacks, significant external forces such as car or motor vehicle accidents, high-impact sports like skiing, and those with fractures caused by underlying pathological conditions to avoid the influence of confounding injuries.^23^ We also excluded patients who did not have enough information on their charts for the calculation of the FRAX score or for the assessment of their falling injury (Figure 1).

**Figure 1.**
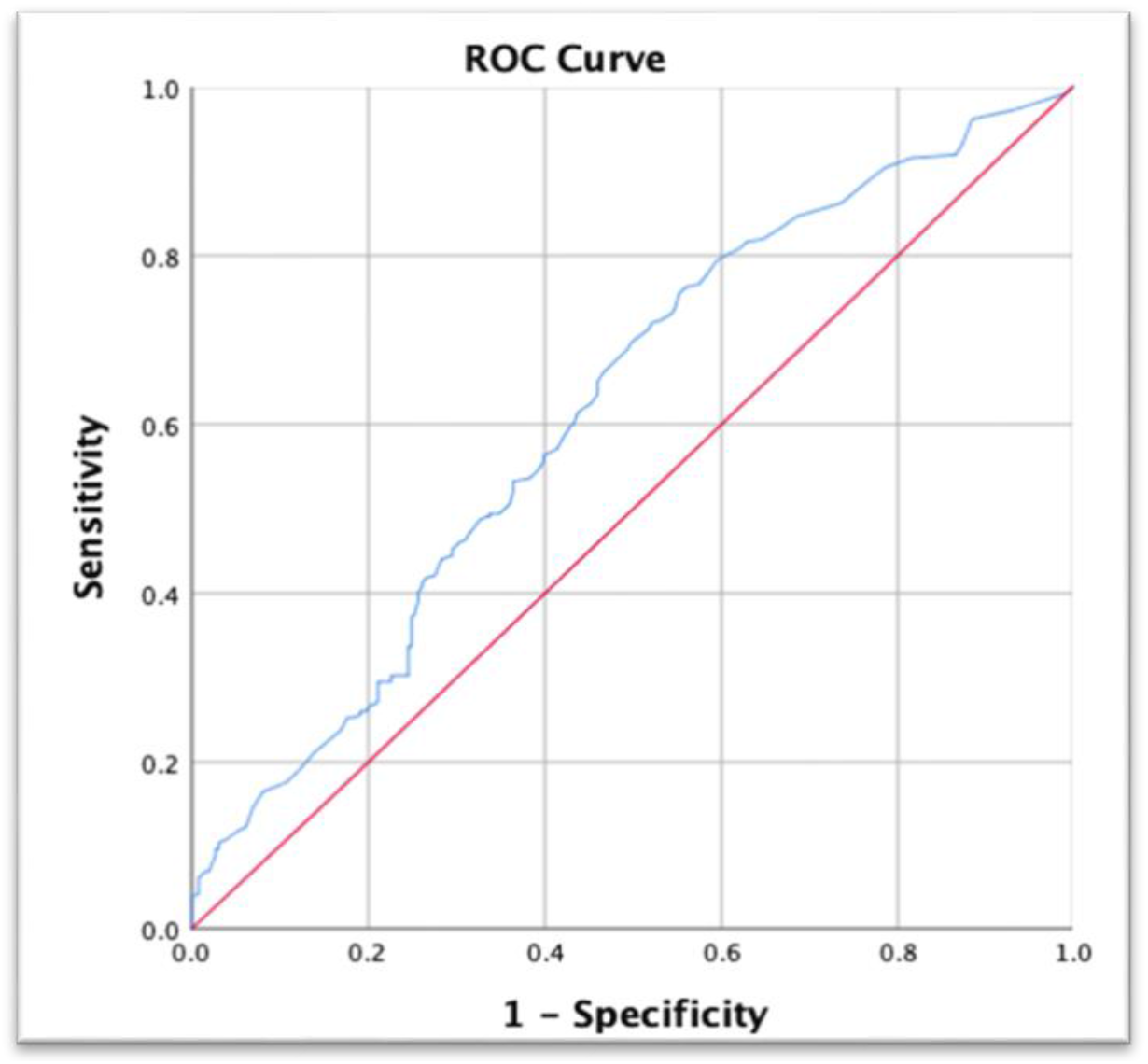
Receiver Operator Characteristics (ROC) Curve for the Original FRAX Model

**Figure 2.**
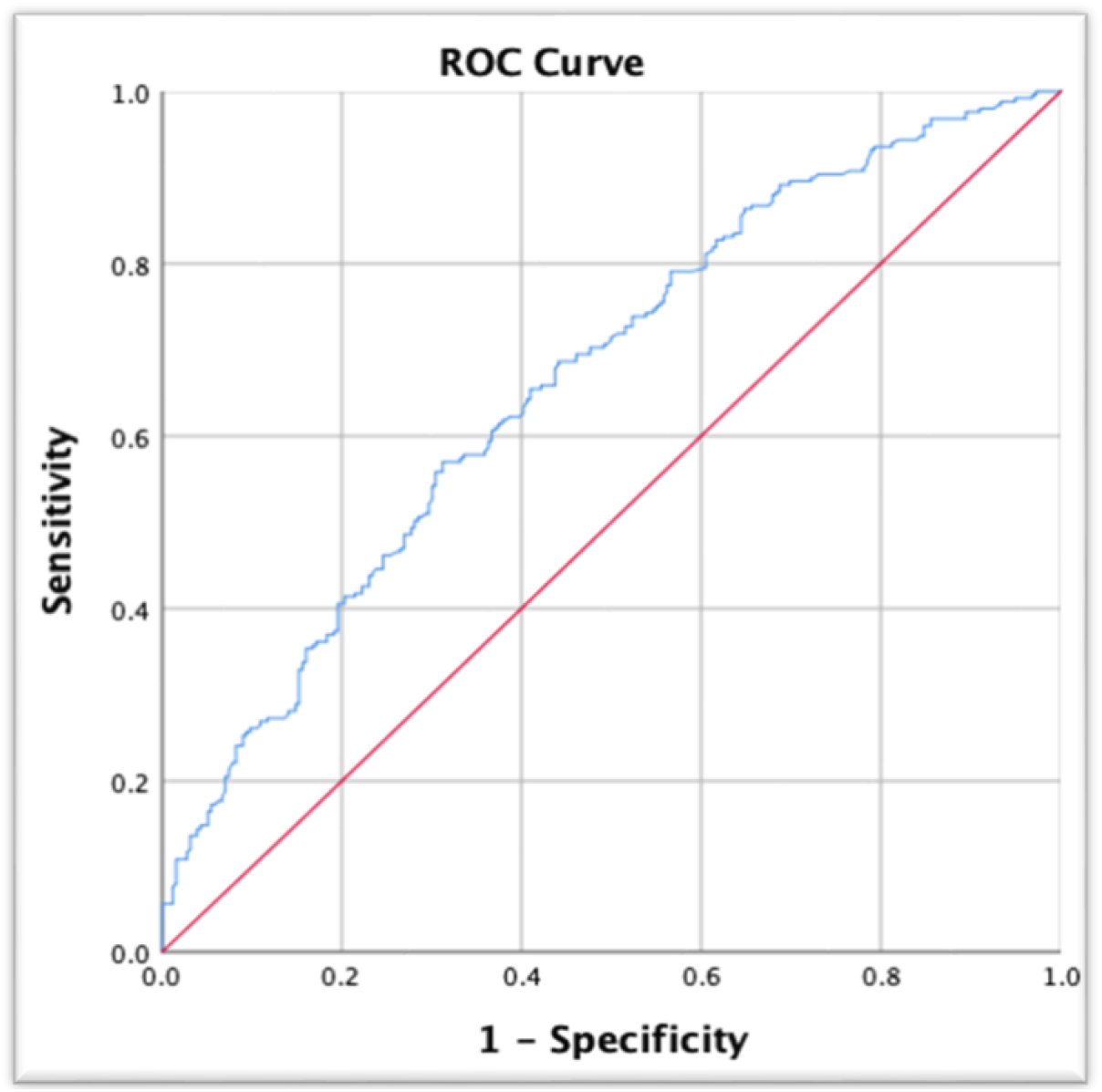
Receiver Operator Characteristics (ROC) Curve for the Best Developed Model. Including the FRAX, Race, Fall Frequency, and Housing Domain of the SVI

### II. Outcomes and Variables

Baseline information of the participants including age, gender, height, weight, body mass index (BMI), race (white, non-white), previous fracture, parent with a fractured hip, smoking status, glucocorticoid use, history of rheumatoid arthritis, secondary osteoporosis, and alcohol consumption. The smoking status was determined at the time of the falling incidence. Glucocorticoids had to be used before or at the time of the fall^24^ To qualify for osteoporosis secondary to a medical condition, the patient had to have one of the following diseases: Type 1 and 2 diabetes, chronic liver disease, inflammatory bowel disease, hypogonadism, celiac disease, premature menopause, cystic fibrosis, renal disease, chronic pancreatitis, sickle cell disease, or hyperthyroidism, clinical visit dates, and notes were collected through the initial query. A thorough search of patients’ electronic health records was done to collect information about the number of previous falls. This included searching for a mention of a fall on all the documented notes, imaging reports, and encounter reasons. The energy of falls was extracted from clinicians’ notes. Low-energy falls were defined as falling from <1 meter and high-energy as >1 meter.^25^ Because most of the time, the falling incidence was described without an explicit mention of the height, it needed to be inferred from the notes by an expert reviewer. As a result, two orthopaedic clinical researchers labeled the patients due to the extracted description of the fall; in case of a disagreement, the opinion of a third experienced observer would help in making the final decision. FRAX scores were calculated using the online calculation tool ([Fracture Risk Assessment Tool](https://frax.shef.ac.uk/FRAX/tool.aspx?country=9)).^24^ The overall SVI and its subcategories, including Socioeconomic Status, Household Characteristics, Racial and Ethnic Minority Status, and Housing Type and Transportation were estimated using the participant’s zip code and CDC’s ATSDR calculation tool ([Agency for Toxic Substances and Disease Registry](https://www.atsdr.cdc.gov/placeandhealth/svi/interactive_map.html)).

### III. Statistical Analysis

Data were analyzed utilizing Statistical Package for the Social Sciences (SPSS Version 28.0, 2021). Descriptive statistics were used to summarize the baseline characteristics of the participants in both groups. The normality of the continuous data was assessed using the Kolmogorov-Smirnov and Shapiro-Wilk tests. T-test, or Mann-Whitney U test, was used to compare normally and non-normally distributed data, respectively. Data were shown as median (Interquartile range; IQR), odds ratio (OR), 95% confidence interval (CI), and percentage (%). A p-value of 0.05 was considered as significant. A binary logistic regression was performed with a stepwise approach to decide on the most relevant predictors based on the level of significance. The performance of the FRAX score, as well as our prediction models, were assessed according to their performance metrics, including the Area Under the Receiver Operating Characteristics Curve (AUC), sensitivity, specificity, positive predictive value (PPV), negative predictive value (NPV), and Youden’s J. The variables from the model with the best performance were chosen to develop the prediction formula. The following formula was used:

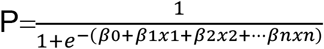

Where “P” is the predicted probability of the outcome; “e” is the base of the natural logarithm (approximately equal to 2.71828); “β0” is the intercept; “β1,β2,…” are the coefficients for the predictor variables; “x1,x2,…” are the predictor variables.

## Results

A total of 261 patients per group were included, and their demographic data are depicted in Table 1. Race, BMI, FRAX, Racial and Ethnic Minority Status, Housing type and Transportation showed significant differences between the hip fracture and no fracture groups.

**Table 1.** Baseline characteristics of the study groups and their comparisons.

| Variable |  | No-fracture Group<br>(n=261) | Fracture Group<br>(n=261) | P Value |
| --- | --- | --- | --- | --- |
| Age (years) |  | 72 (62, 83) * | 74 (67, 80) * | 0.46<sup>‡</sup> |
| Gender | Male | 37.2% | 34.1% | 0.47<sup>†</sup> |
|  | Female | 62.8% | 65.9% |  |
| Race | White | 80.8% | 92.82% | <0.001<sup>†</sup> |
|  | Non-white | 19.14% | 7.17% |  |
| BMI (Kg/m<sup>2</sup>) |  | 26.05 (22.02, 30.34) * | 22.71 (19.55, 26.51) * | <0.001<sup>‡</sup> |
| Energy of falls | High | 16.9% | 11.6% | 0.09<sup>†</sup> |
|  | Low | 83.1% | 88.4% |  |
| Number of falls | None | 44.4% | 50.2% | 0.05<sup>†</sup> |
|  | One | 16.9% | 21.1% |  |
|  | More than one | 38.7% | 28.7% |  |
| FRAX Score |  | 2.50 (0.80, 7.25) * | 4.70 (2.00, 10.00) * | <0.001<sup>‡</sup> |
| SVI (Overall) |  | 0.61 (0.27, 0.79) * | 0.44 (0.27, 0.79) * | 0.40<sup>‡</sup> |
| Socioeconomic |  | 0.36 (0.11, 0.67) * | 0.27 (0.12, 0.67) * | 0.76<sup>‡</sup> |
| Household Characteristics |  | 0.22 (0.07, 0.34) * | 0.22 (0.07, 0.22) * | 0.90<sup>‡</sup> |
| Racial and Ethnic Minority Status |  | 0.69 (0.67, 0.90) * | 0.67 (0.64, 0.90) * | 0.03<sup>‡</sup> |
| Housing Type and Transportation |  | 0.82 (0.82, 0.98) * | 0.82 (0.76, 0.98) * | 0.04<sup>‡</sup> |

\*Median (IQR); †Chi Square; ‡Mann-Whitney U Test

The outcome of the logistic regression test on various factors that showed significant differences between the fracture and no-fracture groups, as well as fall characteristics, including energy and number of falls, are shown in Table 2. The white race also showed a higher rate of fall-induced fracture (OR=2.85; CI:1.56-5.20), although there was a discrepancy between the number of whites and non-whites (86% vs 13%, respectively). Housing Type and Transportation were shown to decrease the chance of experiencing the injury significantly (OR=0.30, CI:0.12-0.80).

**Table 2.**
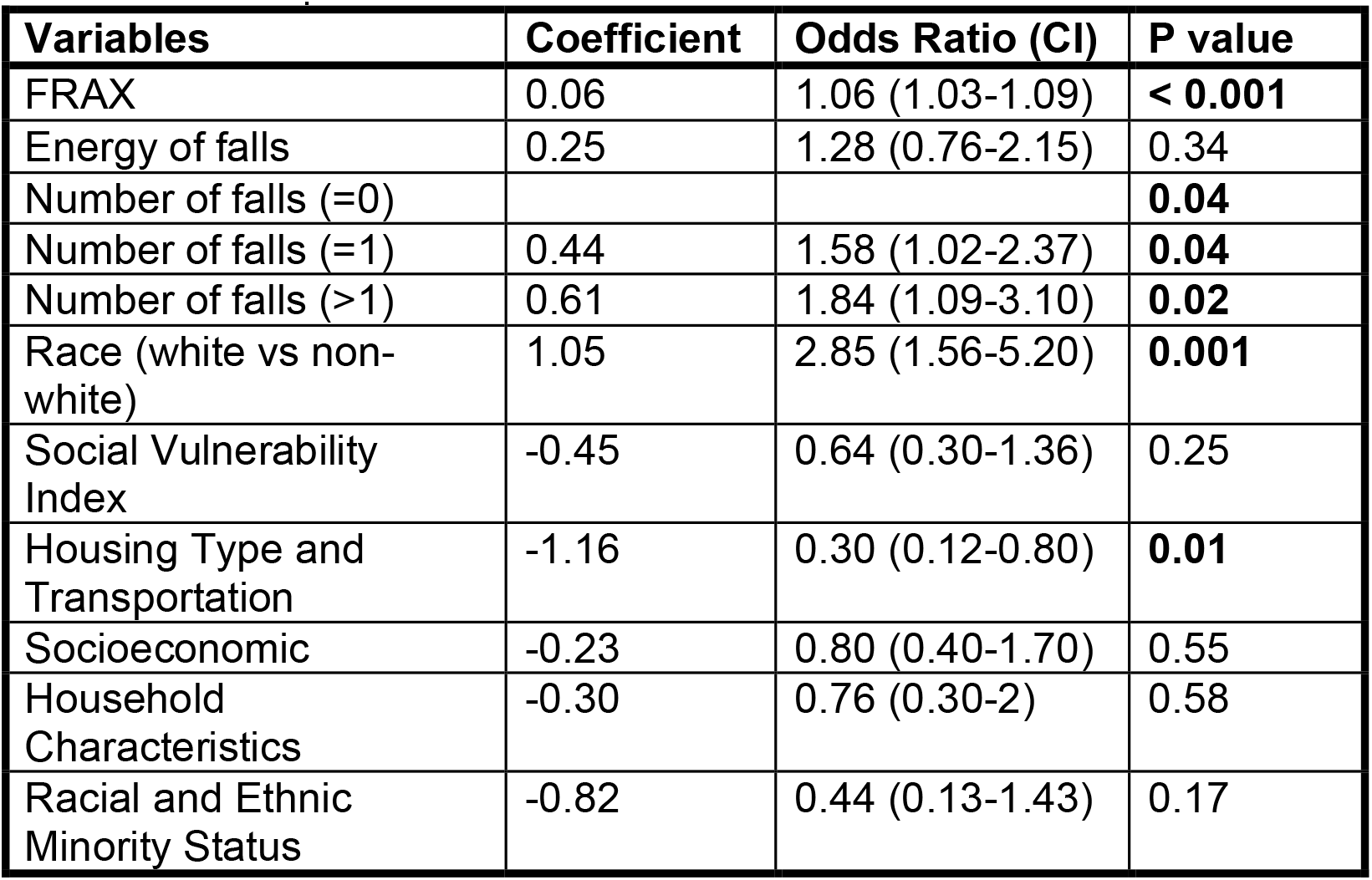
Logistic regression analysis, outlining the impact of different variables on fall-induced hip fracture.

The FRAX tool and the prediction model developed in this study (modified FRAX) were evaluated using the data of the cases and the control group. The performances of the FRAX tool, as well as the modified FRAX, are shown in Table 3. The final prediction model was developed using the coefficients of the best model and deployed on the following web address https://fixus.mgh.harvard.edu/fall/

**Table 3.** The performance metrics across logistic regression models using FRAX tools and the prediction model developed in this study (Modified FRAX).

| **Models** | **AUC** | **Specificity** | **Sensitivity** | **Youden's J** | **NPV** | **PPV** |
| --- | --- | --- | --- | --- | --- | --- |
| FRAX | 62% | 73.6 % | 41.8% | 0.15 | 55.8 % | 61.2% |
| Modified FRAX | 67% | 64.1% | 58.2% | 0.22 | 61.2% | 61.2% |

## Discussions

Almost half of the elderly population experience a fall, and up to 10% have reported a skeletal injury.^3^ A strong association between the propensity to fall and future fracture is mentioned in various studies. These studies showed a greater risk in individuals with a history of multiple falls.^26, 27^ However, the history and frequency of falls have not been included in current prediction models such as the FRAX tool. Falls and hip fractures share many similar risk factors, including increasing age, smoking, alcohol consumption, and frailty; thus, hypothesizing that FRAX and any other prediction model that could predict fractures could also be modified to predict the risk of fall-induced fractures would be reasonable.^12,28^ Including patient-specific characteristics such as race, country of origin, medications, and medical conditions affecting bone quality was considered in several studies to improve the accuracy, validity, and reliability of the FRAX tool^12^ In this study, we investigated the added predictive value of fall characteristics, basic demographics, SVI, and its domains to the FRAX score for the prediction of fall-induced hip fractures. Our results showed that the number of falls, race, Housing type and Transportation subscale of the SVI play a considerable role in predicting the risk of fall-induced hip fractures.

In the nuanced realm of assessing fracture risk, the relationship between fall frequency and the FRAX score emerges as a complex and evolving topic.^12^ In 2015, Harvey et al. conducted a study in a 1,836 male population comparing the predictive value of FRAX and fall frequency for future falls.^27^ They suggested a surprising twist by concluding that the FRAX score is capable of predicting future falls (Hazard Ratio=1.64; CI:1.36-1.97). This finding implied that since falls and fractures share a lot of risk factors, FRAX has an inherent mention of falls. However, this perspective was challenged by subsequent research from the same team in a large-scale study with 7,857 male patients.^14^ Here, they discovered that a patient’s fall frequency indeed has its independent predictive value for future osteoporotic fractures, regardless of their FRAX score. This key insight brought to light the value of including fall history in assessing fracture risk, suggesting that the FRAX score’s effectiveness could be enhanced by integrating this factor.

Our study builds upon this existing body of knowledge, indicating the improved performance of the FRAX score when prior falls are included in the model. While Harvey et al. focused explicitly on male subjects, in the present study, we did not observe any changes when the model was adjusted for sex.^29^ In a study by Kanis et al. involving 21,116 male and female participants, a progressive increase in Hazard Ratios was observed with the rising number of falls.^30^ When these ratios were adjusted for age, sex, and FARX variables, a slight decrease was noted. However, the present study focused specifically on patients with hip fractures caused by falls, providing distinct insights relevant to that population. Moreover, in our study, mainly involving elderly subjects, we did not find the energy of falls to be a significant predictor within the FRAX model.^31, 32^ Notably, we observed a low incidence of high-energy falls among both the cases and controls, aligning with existing literature that low-energy falls are more common in older adults.^33, 34^ Including fall energy in the FRAX model may not significantly affect fracture risk assessment in older adults, but its impact on younger populations warrants further investigation across various age groups.

In predicting fall-induced hip fractures, the significance of incorporating patient sociodemographic characteristics is increasingly recognized. Previous studies have underscored the impact of factors such as race, geographic location, age, economic conditions, and housing on the incidence and management of hip fractures. For instance, disparities in hip fracture rates and outcomes have been observed across different racial and ethnic groups,^35^ while geographic variations highlight the influence of environmental and healthcare access differences.^36^ This understanding has led to the adaptation of the FRAX in various countries, with each version tailored to reflect the specific characteristics and needs of the local population. Our study revealed a notable association between white race and an increased risk of fall-induced injuries, aligning with findings by Ellis et al., who reported that white individuals experienced approximately double the rate of fall-induced hip fractures compared to Hispanic and Black populations and triple the rate observed in Asian/Pacific Islanders.^37^ It is imperative, however, to reflect on the demographic makeup of our study cohort, predominantly composed of white participants, which underscores the need for caution in extrapolating our findings to a broader populations.^38–40^

Whether the broader sociodemographic factors captured by the SVI are of added value in fall-induced injuries is a topic that is not directly addressed in the literature. In this study, we found a significant inverse role for the SVI’s Housing Type and Transportation domain, meaning the less vulnerable individuals had a higher chance of sustaining a fall-induced hip fracture. This counterintuitive result warrants a deeper investigation into the dynamics of the Housing Type and Transportation domain, which includes the number of units in a building, the presence of mobile homes, the number of occupants per room, the presence of a vehicle in a household, and group living arrangements.^41^ One possible explanation could be that a higher number of occupants, typically associated with a higher vulnerability score, might paradoxically confer a protective effect against falls through heightened physical engagement.^42^ Bernhart et al. showed that older adults residing in households with two or more individuals were twice as likely to adhere to aerobic guidelines for physical activity compared to those living in single-adult households.^43^ Similarly, Meghani et al concluded that residing with family plays a crucial role in assisting older adults to stay active and manage their personal responsibilities effectively.^44^ While other socioeconomic factors seem to have a remarkable effect on hip fractures and falls separately, in this study, we did not find a role for other domains of the SVI in fall-induced hip fractures. In contrast, using the Housing-based Index of Socioeconomic Status (HOUSES), Ryu et al. concluded that a higher socioeconomic status reduces the likelihood of falls.^45^ It is important to note, however, that their study included individuals across all age groups. Similarly, a study conducted in the UK highlighted the impact of material deprivation on the incidence of hip fractures among young adults, with a relative risk (RR=1.64; CI:1.57-1.72) in poorer electoral wards.^46^ However, they reported no association in individuals aged 85 and over (RR=0.94, CI:0.87-1.01), suggesting the decreased effect of socioeconomic factors on the elderly, which is consistent with the outcomes of our study.

This study has a few limitations to be considered. While previous studies have shown different results with specific elderly age groups, in this study, because of a relatively small sample size, we could not conduct a group-wise analysis by age. Also, the energy of falls was measured based on a height criterion, and since oftentimes the height was not explicitly mentioned in clinicians’ notes, we relied on the interpretation of the clinical notes made by expert observers. However, if the expert observers were not confident about the interpretation of the fall energy, we would not include that factor for the patient in assessing our prediction models. Despite these limitations, this study is unique in its explicit inclusion of individuals with fall-induced hip fractures. Another strength is the direct extraction of prior fall history from patient records, which reduces the likelihood of recall bias that is often associated with fall questionnaires commonly used in similar studies. Furthermore, this study was the first of its kind, especially in the New England region of the United States, to consider different socioeconomic and demographic characteristics of patients and their relevance to fall-induced injuries.

## Conclusion

This study highlighted the importance of the history and frequency of prior falls for the risk assessment of fall-induced hip fractures. Our results revealed the role of the Housing and Transportation subscale of the SVI, which also happens to be a modifiable risk factor. Adding these patient-specific factors to the widely used and accepted FRAX model has led to a modified FRAX that outperformed the FRAX tool in the prediction of fall-induced hip fractures. However, there is still potential for further improvement in the FRAX model. We recommend future research focus on age group stratification and validation of the impact of social determinants of health and socioeconomic characteristics of patients. This should include, but not be limited to, factors like the SVI and the Area Deprivation Index, as well as the characteristics of the falls themselves. These prediction models should be built using large and granular data and should be externally validated to achieve higher validity and generalizability.

## Data Availability

All data produced in the present study are available upon reasonable request to the authors

## Acknowledgment

The authors would like to acknowledge the patients and healthcare professionals whose data and experiences form the backbone of this study, contributing significantly to our understanding of fall-induced injuries.

## Notes

### Competing Interest Statement

The authors have declared no competing interest.

### Funding Statement

This study did not receive any funding

### Author Declarations

Institutional Reveiw Board of Massachusetts General Hospital gave ethical approval for this work

